# Measurement strategy alters inferred age-dependent accumulation and mortality risk of mosaic Y loss

**DOI:** 10.64898/2026.03.09.26347951

**Authors:** Akshay Ware, Michael Weyrich, Sameen Fatima, Tenglong Xu, Srisurekha Radhakrishnan, Paul Kapfer, Xiao Yang, Lili Schiethe, Lukas Zanders, Sebastian Cremer, Silvia Mas-Peiro, Stefanie Dimmeler, Timotheus Speer, Andreas M. Zeiher, Wesley T. Abplanalp

**Author notes:** corresponding author, Address for correspondence: Wesley T. Abplanalp, Prof. of Medicine Goethe University Theodor-Stern-Kai 7, 60590 Frankfurt Germany. contributed equally (Andreas M. Zeiher, Wesley T. Abplanalp).

## Abstract

Mosaic loss of Y chromosome (mLOY) is a widely used biomarker of biological aging, yet whether its inferred age-dependent accumulation and associated clinical risk are invariant to measurement strategy remains unclear. We compared intensity-based and phase-based quantification approaches in 223,251 men from the UK Biobank to determine how analytic definitions influence estimates of mLOY burden, risk thresholds and population prevalence. Phase-based quantification revealed a steeper and more stable age-dependent accumulation of mLOY and identified excess mortality risk at lower mosaic burdens than intensity-based metrics. These differences shifted the inferred onset of biological risk and expanded the proportion of individuals classified as affected from 5.3% to 19.2%. Conventional thresholding preferentially excluded low-burden mosaicism, compressing risk gradients and reducing statistical resolution for downstream associations. These findings show that analytic definitions materially alter inferred accumulation dynamics, risk thresholds and population prevalence of mosaic Y loss.

## Introduction

Mosaic loss of the Y chromosome (mLOY) is one of the most prevalent somatic alterations observed in aging men and is increasingly interpreted as a biomarker of biological aging.^1,2^ Its reported associations with cardiovascular disease, cancer, immune dysfunction and mortality have positioned mLOY as a putative contributor to age-related pathology rather than merely a correlate of aging.^1–6^ However, these biological inferences rely on quantitative definitions of mosaic burden that vary substantially across analytic pipelines, raising the question of whether the apparent age-dependent accumulation and clinical risk landscape of mLOY are invariant to how the biomarker is measured.

Several molecular approaches are available to detect mLOY, including digital PCR, whole-genome and whole-exome sequencing, and genotyping array based methods.^1,2^ Among these, genotyping arrays are most widely used in large population studies due to their scalability, cost-effectiveness, and availability in resources such as the UK Biobank.^3^ Array-based detection infers mLOY from deviations in probe intensity or allelic balance across the Y chromosome, serving as proxies for the fraction of leukocytes lacking chromosome Y.

A commonly used array-based metric is the median log R ratio of the Y chromosome (mLRRY), which summarizes normalized probe intensity across the male-specific region after exclusion of pseudoautosomal regions (PARs).^4^ Increasingly negative mLRRY values reflect progressive loss of Y-bearing cells, and predefined thresholds are used to classify individuals as mLOY-positive.^2,3^ However, because mLRRY relies on global intensity shifts, it is susceptible to technical artifacts and reduced sensitivity at low clonal fractions, limiting its ability to detect low-frequency mosaicism.

More recently, phase-based algorithms such as MoChA enabled more precise detection of mosaic chromosomal events using phased genotype data.^7–9^ MoChA detects mLOY using allelic imbalance at heterozygous variants in pseudoautosomal region 1 (PAR1), incorporating phased B-allele frequency and log R ratio information across sex chromosome specific regions. This approach allows for sensitive detection of sex chromosome mosaicism and quantitative estimation of clonal cell fractions, substantially improving specificity and dynamic range compared with intensity-based methods.^6^

Regardless of these advantages, MoChA requires high-quality phased genotypes and sufficient heterozygosity in PAR1, which may not be uniformly available across cohorts or array platforms.^10^ In addition, its computational and preprocessing demands may limit adoption in some analytic pipelines. Consequently, both mLRRY and MoChA continue to be used in parallel in the literature, often without direct comparison or harmonization of results.

Despite widespread use of mLOY as an aging biomarker, it remains unclear whether key inferences about accumulation, risk onset and population prevalence depend on how mosaic burden is quantified.^11^ In large-scale studies, somatic alterations are often summarized using continuous metrics or dichotomized using conservative thresholds to reduce technical noise. However, these methodological choices can substantially alter estimates of prevalence, age dependence, and disease associations. Consequently, differences in reported aging patterns across studies may reflect measurement strategy as much as underlying biology. Clarifying the impact of measurement choices is therefore essential for accurate interpretation of population-scale somatic aging.

Here, we systematically evaluate whether the inferred accumulation dynamics and mortality risk associated with mLOY depend on the analytic definition of mosaic burden. Using 223,251 male UK Biobank participants, we compare intensity-based and phase-based quantification to determine how measurement strategy alters estimates of age-dependent burden, risk thresholds, and population prevalence of biologically meaningful mosaicism. We quantified overlap and discordance between methods, evaluated age- and smoking-associated trends, and assessed agreement in inferred clonal fractions. We further examined technical variability and specificity at low mosaic burdens. Finally, we tested whether methodological differences produce divergent associations with all-cause mortality, linking quantification strategy to epidemiologic interpretation.

## Results

### Overall Statistics and Age-Associated Trends in %mLOY Detection Differ Between Methods

Using MoChA, %mLOY was identified in 44,606 male participants (19.98%) from the UK Biobank, whereas the mLRRY approach classified 110,486 individuals (49.48%) as mLOY-positive (Table 1). This higher prevalence for mLRRY was driven primarily by additional low-burden calls, consistent with increased sensitivity to intensity noise at small clonal fractions. Stratification of mLOY-positive individuals by estimated mosaic fraction revealed substantial method-dependent differences at low mLOY burden. In the lowest mLOY bin (0–5%), mLRRY classified 78,579 men (35.19% of the cohort) as mLOY-positive, compared with only 12,823 men (5.74%) identified by MoChA. In contrast, the two methods showed close agreement at moderate mosaic fractions, with similar proportions of individuals classified in the 5-10% and 10-15% mLOY bins. Above approximately 20% mLOY, prevalence estimates were nearly identical across methods, indicating strong concordance at higher mosaic burden.

**Table 1.** Distribution of mosaic loss of chromosome Y (mLOY) burden in male UK Biobank participants detected by MoChA and mLRRY.

| %mLOY | Total mLOY (per method) |  | Common mLOY (overlap) |  |
| --- | --- | --- | --- | --- |
|  | MoChA (Count %)<br>(N=44,606) | mLRRY (Count %)<br>(N=110,486) | MoChA (Count %)<br>(N=35,845) | mLRRY (Count %)<br>(N=35,845) |
| 0-5 | 12823 (5.74) | 78579 (35.19) | 7048 (3.15) | 14690 (6.58) |
| 5-10 | 18246 (8.17) | 19814 (8.87) | 15357 (6.87) | 10457 (4.68) |
| 10-15 | 5929 (2.65) | 5386 (2.41) | 5855 (2.62) | 4611 (2.06) |
| 15-20 | 2817 (1.26) | 2456 (1.10) | 2808 (1.25) | 2183 (0.97) |
| 20-25 | 1480 (0.66) | 1417 (0.63) | 1480 (0.66) | 1268 (0.56) |
| 25-30 | 935 (0.41) | 856 (0.38) | 934 (0.41) | 784 (0.35) |
| 30-35 | 697 (0.31) | 575 (0.25) | 695 (0.31) | 562 (0.25) |
| 35-40 | 504 (0.22) | 371 (0.16) | 501 (0.22) | 359 (0.16) |
| 40-45 | 361 (0.16) | 289 (0.12) | 359 (0.16) | 285 (0.12) |
| 45-50 | 311 (0.13) | 251 (0.11) | 309 (0.13) | 248 (0.11) |
| 50-55 | 236 (0.10) | 152 (0.06) | 235 (0.10) | 151 (0.06) |
| 55-60 | 167 (0.07) | 112 (0.05) | 165 (0.073) | 110 (0.04) |
| 60-65 | 98 (0.04) | 94 (0.04) | 98 (0.043) | 89 (0.03) |
| 65-70 | 2 (9e-04) | 61 (0.02) | 1 (4e-04) | 41 (0.018) |
| 70-75 | 0 (0) | 44 (0.01) | 0 (0) | 5 (0.0022) |
| 75-80 | 0 (0) | 22 (0.0099) | 0 (0) | 0 (0) |
| 80-85 | 0 (0) | 5 (0.0022) | 0 (0) | 1 (4e-04) |
| 85-90 | 0 (0) | 1 (4e-04) | 0 (0) | 1 (4e-04) |
| 90-95 | 0 (0) | 1 (4e-04) | 0 (0) | 0 (0) |

**mLOY:** mosaic loss of Y chromosome **MoChA:** mosaic chromosomal alteration (pipeline) **mLRRY:** median log-R ratio of the Y chromosome (pipeline) **Common:** overlapping subset of participants classified as mLOY-positive by both MoChA and mLRRY.

Differences between methods were primarily driven by participants in older age groups and those with low estimated mosaic cell fractions. Among men aged ≥65 years, MoChA detected mLOY in 38.0% of participants, while mLRRY identified 62.5%, indicating method-dependent sensitivity at later ages (Table 2, Supplemental Table 1).

**Table 2.** Age-stratified prevalence of mLOY among male UK Biobank participants assessed using MoChA and mLRRY.

|  | MoChA |  | mLRRY |  |
| --- | --- | --- | --- | --- |
| Age | Total<br>(N=223,251) | mLOY (Count %)<br>(N=44,406) | Total<br>(N=222,974) | mLOY (Count %)<br>(N=110,486) |
| <41 | 2552 | 70 (2.74) | 2543 | 827 (32.52) |
| 41-42 | 10105 | 274 (2.71) | 10084 | 3348 (33.20) |
| 43-44 | 10663 | 359 (3.36) | 10650 | 3636 (34.14) |
| 45-46 | 10970 | 432 (3.93) | 10950 | 3937 (35.95) |
| 47-48 | 11479 | 558 (4.86) | 11457 | 4242 (37.02) |
| 49-50 | 12131 | 810 (6.67) | 12108 | 4754 (39.26) |
| 51-52 | 12675 | 1162 (9.16) | 12660 | 5201 (41.08) |
| 53-54 | 13399 | 1604 (11.97) | 13392 | 5927 (44.25) |
| 55-56 | 14678 | 2174 (14.81) | 14661 | 6830 (46.58) |
| 57-58 | 15658 | 2945 (18.80) | 15642 | 7859 (50.24) |
| 59-60 | 19325 | 4368 (22.60) | 19304 | 10226 (52.97) |
| 61-62 | 22691 | 5979 (26.34) | 22666 | 12555 (55.39) |
| 63-64 | 20688 | 6287 (30.38) | 20661 | 11904 (57.61) |
| 65-66 | 20294 | 7068 (34.8) | 20285 | 12375 (61.00) |
| 67-68 | 16929 | 6716 (39.67) | 16910 | 10845 (64.13) |
| 69-73 | 9014 | 3800 (42.15) | 9001 | 6020 (66.88) |

**mLOY:** mosaic loss of Y chromosome **MoChA:** mosaic chromosomal alteration (pipeline) **mLRRY:** median log-R ratio of the Y chromosome (pipeline)

Both methods demonstrated a strong positive association between age and mLOY burden. Linear regression of age against estimated %mLOY revealed robust age-dependent increases for both approaches, with a significantly steeper increase observed using MoChA (β = 3.73% per decade, 95% CI 3.59–3.88) compared with mLRRY (β = 2.31% per decade, 95% CI 2.25–2.36) both p < 2.2 × 10^-16^ (Figure 1A). Despite broadly concordant age-related trends, these measurement strategies yield qualitatively distinct age-dependent accumulation patterns of mLOY burden, indicating that the apparent pace and variability of this aging biomarker depend on analytic definition. Notably, MoChA exhibited reduced residual variance and narrower confidence intervals around fitted estimates, suggesting greater stability of age-associated estimates. This likely reflects MoChA’s reliance on phased B-allele frequency deviations in PAR1, which are less susceptible to global intensity noise than logR ratio-based metrics.

**Figure 1.**
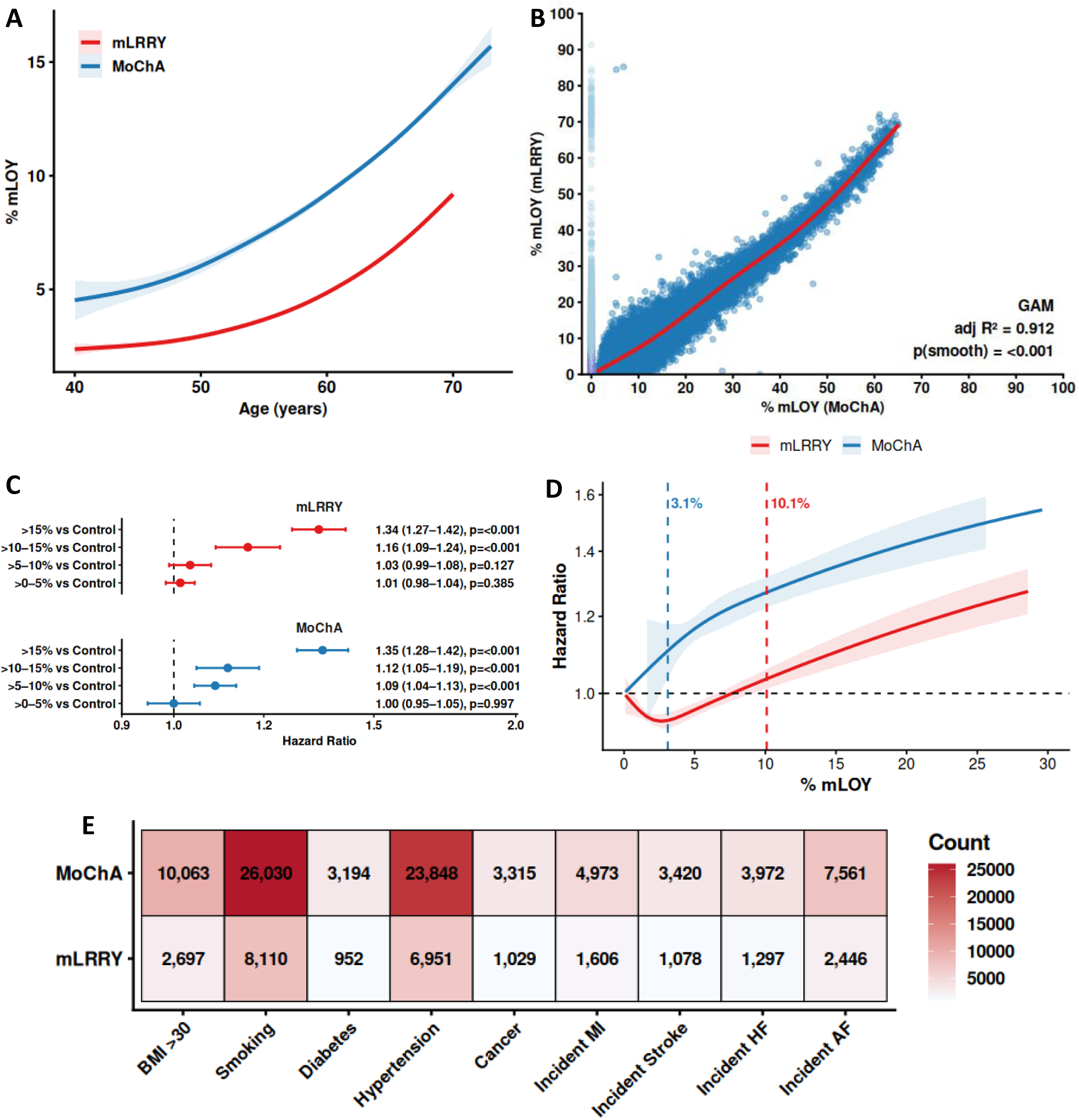
mLOY measurement, concordance, and association with mortality across methods. **(A)** Generalized additive model (GAM) spline curves showing the association between age and percentage mLOY. Shaded areas represent 95% confidence intervals. **(B)** Concordance between MoChA- and mLRRY-derived mLOY estimates in paired samples. The GAM fit demonstrates strong agreement between methods (adjusted R² = 0.91, *P* < 0.001 for smooth term). **(C)** Association of mLOY categories with all-cause mortality. Hazard ratios (HRs) are estimated from Cox proportional hazards models comparing mLOY categories (>0-5%, >5-10%, >10-15%, >15%) to controls (0%), adjusted for age, body mass index, smoking status, alcohol consumption, and diabetes. Points indicate HRs and error bars denote 95% confidence intervals. **(D)** Dose-response relationship between mLOY and all-cause mortality estimated using Cox models with natural cubic splines. HRs are shown relative to 0% mLOY, with shaded 95% confidence intervals. Vertical dashed lines indicate spline-derived thresholds at which the lower confidence interval exceeds unity (MoChA: 3.1%; mLRRY: 10.1%). **(E)** Heatmap of selected demographic, lifestyle, and clinical characteristics among individuals exceeding method-specific mLOY thresholds. Values represent counts and percentages relative to all individuals with detectable mLOY within each method.

### Inter-method Concordance of mLOY Estimates (MoChA vs mLRRY)

MoChA and mLRRY derived estimates of mLOY were strongly correlated in paired samples (GAM spline adjusted R² = 0.912, p for smooth < 0.001; Figure 1B), indicating substantial shared signal between methods. However, the relationship exhibited modest non-linearity at lower mLOY levels, consistent with systematic method-dependent divergence, particularly at low mosaic fractions. These findings indicate that while both approaches capture overlapping biological information, technical and algorithmic differences meaningfully influence inferred mLOY burden, supporting the conclusion that measurement strategy contributes to observed heterogeneity in aging trajectories.

### Method discordance is concentrated at low mosaic burden

Across both pipelines, 119,247 unique participants were classified as mLOY positive by at least one method. Of these, 35,845 individuals were concordantly identified by both MoChA and mLRRY, while the remainder were method-specific (Supplemental Figure 1A). Discordant calls were overwhelmingly concentrated at low mosaic burdens, demonstrating that the definition of low-level mosaicism critically determines how early clonal loss is inferred to emerge during aging. Rank-ordered %mLOY plots for method-specific calls showed that most discordant detections occurred in the 0–5% range, consistent with a regime in which array-based mLOY estimates are most susceptible to technical noise.

Among the 35,845 participants with concordant mLOY calls, MoChA and mLRRY showed similar overall burden distributions, with most events occurring at low clonal fractions (Table 1). However, mLRRY assigned a greater proportion of individuals to the lowest bin (0–5%), whereas MoChA classified relatively more individuals in the 5–10% range. Above 10% mLOY, bin-wise frequencies were closely aligned between methods, indicating strong concordance at moderate-to-high mosaic burdens.

Direct comparison of %mLOY estimates further demonstrated that inter-method disagreement was greatest at low burden and decreased progressively with increasing mosaicism. Among participants classified as mLOY-positive by both methods, relative variability (coefficient of variation, SD/mean) was highest in the 0–5% bin (median 0.386; IQR 0.173–0.718; n = 7,048) and declined steadily at higher burdens (e.g., 0.212 at 11–15%, 0.143 at 16–20%, and 0.025 at 56–60%) (Supplemental Figure 1B). Overall, relative variability showed a strong inverse association with mLOY burden (Spearman’s ρ = −0.40, p < 2.2 × 10^-1^□), indicating increasing concordance as biological signal strength increased.

Consistent with these findings, changes in mLOY categorization were primarily limited to the low-burden regime. At low mosaic fractions, mLRRY detected significantly more mLOY-positive people than MoChA, but classification frequencies converged rapidly with rising %mLOY, leading in strong agreement at moderate and higher loads. This pattern indicates that MoChA applies more conservative calling at very low mosaic fractions, preferentially retaining events with stronger signal-to-noise and thus greater biological relevance. These low-burden discrepancies prompted additional analyses to investigate how mLRRY thresholding affects cohort makeup and overlaps with MoChA derived mLOY calls.

### Impact of Conventional mLRRY Thresholding on Sample Retention and Method Concordance

Prior UK Biobank studies have often applied a conservative mLRRY threshold to limit false-positive calls arising from array intensity noise. An empirically derived cutoff has been used to dichotomize individuals as mLOY-positive or negative, prioritizing specificity over sensitivity to low-fraction mosaicism.^2^ To place our continuous mLRRY and MoChA analyses in this commonly used framework, we examined how applying this threshold (corresponding ∼8.81% mLOY) alters cohort composition and concordance with MoChA-detected events. The derivation of this threshold is shown in Supplemental Figure 2.

The 8.81% threshold eliminated 96,103 men who were identified as mLOY positive using continuous mLRRY estimations and/or MoChA. Exclusions were mostly centered in the low mosaicism range (<10%), when MoChA detected organized and age associated mLOY signals. As a result, conventional mLRRY thresholding significantly reduces effective sample size while selectively excluding low-fraction mosaic events, which directly contributes to the observed disparities in mLOY prevalence and burden distributions between thresholded mLRRY and MoChA based analyses.

### Association Between Method-Dependent mLOY Burden and All-cause Mortality

Using predefined mLOY burden ranges, MoChA derived estimates demonstrated a clear and graded association with all-cause mortality. Relative to individuals without detectable mLOY (0%), no excess risk was observed at very low clonal burdens (>0–5%; HR = 1.00, 95% CI 0.95–1.05). However, mortality risk increased progressively across higher ranges, with modest elevations at >5–10% (HR = 1.09, 95% CI 1.04–1.13; p = 1.1 × 10^⁻^□) and >10–15% (HR = 1.12, 95% CI 1.05–1.19; p = 7.3 × 10^⁻^□), and a pronounced increase at >15% mLOY (HR = 1.35, 95% CI 1.28–1.42; p = 5.1 × 10^⁻³^□) (Figure 1C, Supplemental Figure 1C). These findings indicate that the observed dose–response relationship between mLOY and mortality varies according to quantification strategy, with phase-based methods revealing a graded risk gradient across lower mosaic burdens.

In contrast, mLRRY derived estimates exhibited a less consistent pattern at lower clonal burdens. No significant association with mortality was observed in the >0–5% (HR = 1.01, 95% CI 0.98–1.04; p = 0.39) or >5–10% ranges (HR = 1.03, 95% CI0.99–1.08; p = 0.13). A clearer increase in risk emerged only at higher mLOY levels, with significant associations observed at >10–15% (HR = 1.16, 95% CI 1.09–1.24; p = 6.5 × 10⁻□) and >15% (HR = 1.34, 95% CI 1.27–1.42; p = 2.3 × 10⁻ ²□). This suggests reduced sensitivity of intensity-based measures in detecting risk gradients at low-to-intermediate clonal burdens.

When restricting the mLRRY analysis to individuals exceeding the conventional 8.81% threshold (n = 14,379), a more coherent dose response relationship became apparent. Within this subset, mortality risk increased with higher mLOY burden, reaching statistical significance at >10–15% (HR = 1.17, 95% CI 1.10–1.25; p = 2.0 × 10⁻□) and >15% (HR = 1.36, 95% CI 1.28–1.43; p = 2.7 × 10⁻²□). Associations at lower ranges were attenuated and not statistically significant, consistent with reduced power and the exclusion of low-burden clones.

Together, these analyses show that estimated risk gradients differ depending on how mosaic burden is quantified. Phase-based MoChA captures a continuous and graded risk increase across the full spectrum of mLOY, whereas intensity-based mLRRY primarily detects associations at higher clonal burdens unless thresholding is applied. These findings highlight the importance of measurement strategy in defining both the sensitivity and functional form of mLOY-mortality associations in population-scale studies.

### Method Dependent Risk Thresholds and Population Capture

To directly compare how mLOY quantification strategy influences risk estimation, we modelled MoChA and mLRRY derived mLOY as continuous exposures using spline-based Cox regression. The two methods yielded qualitatively distinct risk functions.

MoChA demonstrated a gradual and continuous increase in mortality risk across the mLOY spectrum, with hazard ratios exceeding unity at relatively low clonal fractions (∼3.1% mLOY; HR ≈ 1.11, 95% CI 1.04–1.18) (Figure 1D). In contrast, mLRRY exhibited a delayed and compressed risk profile, with risk becoming detectable only at substantially higher levels (∼10.1% mLOY; HR ≈ 1.03, 95% CI 1.01–1.06). These differences indicate that phase-based inference captures biologically relevant signal at lower mosaic fractions, whereas intensity-based estimates require higher burden to achieve comparable statistical resolution.

Applying these empirically derived thresholds revealed marked divergence in the fraction of individuals classified as having elevated mLOY. The MoChA defined threshold (≥3.1%) identified 42,906 individuals (19.2% of all males), whereas the mLRRY threshold (≥10.1%) identified only 11,928 individuals (5.3%). Thus, the choice of measurement approach results in a ∼4-fold difference in the size of the high-mLOY population.

We next assessed how these method-specific thresholds map onto clinical characteristics using absolute counts. Individuals exceeding the MoChA threshold showed substantial enrichment for established risk factors, including smoking (n = 26,030), obesity (n = 10,063), diabetes (n = 3,194), and hypertension (n = 23,848), along with a higher burden of incident cardiovascular outcomes, including myocardial infarction (n = 4,973), stroke (n = 3,420), heart failure (n = 3,972), and atrial fibrillation (n = 7,561). In contrast, individuals exceeding the mLRRY threshold represented a more stringently selected subset, with consistently lower counts across these traits, including smoking (n = 8,110), obesity (n = 2,697), diabetes (n = 952), and hypertension (n = 6,951), as well as fewer incident cardiovascular events (myocardial infarction: n = 1,606; stroke: n = 1,078; heart failure: n = 1,297; atrial fibrillation: n = 2,446) (Figure 1E).

Together, these findings demonstrate that mLOY outcome associations are not invariant to measurement strategy. Instead, the choice of method shifts both the effective risk threshold and the population subset captured, thereby altering the inferred relationship between mosaic chromosomal alterations and clinical outcomes. Phase based approaches (MoChA) provide a more sensitive and graded representation of clonal burden, whereas intensity-based measures (mLRRY) preferentially detect higher-burden clones and compress the observable risk landscape.

### Smoking, Alcohol, Diabetes, and Age Stratify mLOY Burden Across Detection Pipelines

Stratified analyses revealed broadly consistent demographic patterns of mLOY across both detection pipelines. Smoking status was the strongest lifestyle correlate of mLOY. Smokers (current or former) exhibited a significantly higher mLOY burden than non-smokers in both methods. Median %mLOY was higher among smokers compared with non-smokers for MoChA (7.37% vs. 6.24%; p < 2.2 × 10^-16^, Wilcoxon rank-sum test) and mLRRY (3.20% vs. 2.64%; p < 2.2 × 10^-16^), indicating robust and method independent effects of smoking on mLOY burden (Supplemental Figure 1D).

Alcohol consumption showed no meaningful association with mLOY burden in either pipeline. Median %mLOY was similar between alcohol consumers and non-consumers for MoChA (6.86% vs. 6.95%; p = 0.36) and mLRRY (2.92% vs. 2.90%; p = 0.83), suggesting minimal influence of alcohol intake on detectable mLOY burden (Supplemental Figure 1E). Diabetes status was associated with modest but statistically significant differences in mLOY burden. Individuals with diabetes exhibited slightly higher median %mLOY than those without diabetes for both MoChA (7.18% vs. 6.85%; p = 3.0 × 10^-5^) and mLRRY (3.03% vs. 2.92%; p = 3.8 × 10^-4^) (Supplemental Figure 1F). However, the magnitude of these differences was small compared with smoking-related effects, indicating a weaker and potentially indirect association. BMI showed little evidence of association with mLOY burden across both detection pipelines. Mean BMI was similar for MoChA and mLRRY (27.6 vs. 27.7 kg/m²), with comparable distributions across BMI categories (MoChA: normal 11,502, overweight 22,446, obese 10,482; mLRRY: normal 27,834, overweight 53,703, obese 26,249), suggesting that adiposity is not a major determinant of detectable mLOY burden (Supplemental Figure 1G).

Across all strata, age remained a dominant determinant of mLOY burden. Within each smoking, alcohol, and diabetes category, %mLOY increased monotonically with age for both methods, consistent with age-dependent clonal hematopoiesis. Together, these findings demonstrate that MoChA and mLRRY capture concordant biological signals for major demographic correlates of mLOY, while differing in sensitivity at lower mosaic burdens.

## Discussion

Our findings demonstrate that the apparent accumulation pattern and clinical risk landscape of a canonical aging biomarker are not intrinsic properties of biology alone, but are partly determined by how mosaic burden is quantified.^11^ We found substantial discrepancies in the number and distribution of mLOY calls, particularly in the low mosaicism regime, with only 35,845 individuals concordantly classified as mLOY positive by both methods.

Although focused on mLOY, these results have broader implications for the interpretation of somatic aging biomarkers, including clonal hematopoiesis and mosaic chromosomal alterations, where analytic thresholds and quantification strategies are frequently treated as neutral technical choices rather than determinants of biological inference. Similar measurement choices arise in studies of clonal hematopoiesis, mosaic chromosomal alterations, and other blood-based aging biomarkers, where thresholds and summary metrics are frequently applied to noisy signals.^11,12^ Our results highlight the need to distinguish biological aging processes from artifacts introduced by measurement strategy, particularly in large population cohorts where small biases can have substantial effects on inferred aging trajectories. More broadly, analytic choices can materially influence how somatic mosaicism is interpreted in population studies.

Depending on whether mLOY is quantified using continuous intensity metrics, conservative thresholds, or phase-based models, estimates of prevalence, age dependence, and mortality risk can differ markedly. This suggests that some apparent features of biological aging in epidemiologic studies may reflect measurement choices rather than underlying biology alone.

Importantly, many previously published UK Biobank analyses of mLOY have implicitly conditioned on the application of a conservative mLRRY threshold,commonly corresponding to ∼8-9% mLOY^5,6^.^3,4^ Our findings reveal that this practice systematically excludes over 96,103 men, primarily those with low-fraction mosaicism, thereby potentially introducing selection bias toward higher-burden events in epidemiologic analyses. As a result, prior estimates of mLOY prevalence, age trajectories, and disease associations may have been skewed toward individuals with higher-burden mosaic events.

By expanding detection of low-burden clones that exhibit graded risk associations, phase-based quantification increases the statistical resolution to detect downstream clinical associations. Thus, the apparent gain in power is not merely a technical advantage but reflects a redefinition of which mosaic burdens are considered biologically relevant.

The mortality analyses further demonstrate that measurement choice directly shapes biological inference. MoChA based ranges revealed a graded relationship between increasing mLOY burden and all-cause mortality, whereas continuous mLRRY produced non-monotonic risk estimates in intermediate quintiles, likely reflecting residual technical noise at low mosaic fractions. Thresholded mLRRY yielded patterns more similar to MoChA but with reduced statistical power due to sample attrition. Together, these results indicate that phase-based approaches may provide more stable risk estimation across the mLOY spectrum, particularly at lower burdens.

A central implication is that measurement strategy shifts the estimated onset of observable risk. Phase-based quantification revealed excess mortality risk at substantially lower mosaic burdens than intensity-based metrics, indicating that the apparent timing at which clonal loss becomes clinically relevant depends on analytic definition. Consequently, some reported thresholds for biologically meaningful mLOY may reflect measurement conventions rather than discrete biological transitions.

These differences have meaningful implications for downstream genetic and phenotypic studies. Although MoChA is algorithmically sensitive to low-level mosaicism, its probabilistic modeling and quality-filtering framework yields a more conservative call set at very low estimated mLOY fractions. MoChA appears to offer more stringent and possibly more specific calls, particularly at mLOY <5%, which may reduce false positives. However, this comes at the cost of reduced sensitivity and the need for high-quality phasing and sufficient heterozygous markers in PAR1. Conversely, mLRRY provides broader coverage but with higher variability and potential overcalling in borderline cases.

In light of these differences, several practical considerations for study design emerge. Continuous mLRRY is well suited for population-level descriptions, but becomes increasingly noisy at low mLOY fractions. In this setting, applying a conservative threshold (≈8–10%) or modeling mLRRY in bins can improve interpretability, although such thresholds inevitably exclude a large group of individuals with low-level mosaicism and may introduce selection bias.

For association analyses, phase-based approaches such as MoChA provide more robust inference. By leveraging haplotype information and stricter quality control, MoChA yields more stable and biologically consistent dose–response relationships, particularly at lower mosaic fractions. Increasing stringency further reduces call numbers primarily among borderline events but improves specificity and typically strengthens effect estimates, reflecting a trade-off between sensitivity and reliability.

In practice, mLRRY is suited for descriptive population-level detection, whereas MoChA provides higher-confidence burden estimates for association and risk analyses. Reporting results across multiple parameterizations may help ensure that conclusions are not driven by measurement strategy alone.

Our findings align with recent frameworks in CHIP mutation curation, such as those by Vlasschaert et al., which emphasize cautious interpretation of low-VAF events.^12^ Applying similar logic to mLOY detection, we recommend stratifying mLOY burden into biologically meaningful bins and considering method-specific thresholds when integrating mLOY calls into association studies.

Collectively, these findings indicate that the biological interpretation of mosaic Y loss, its apparent age-dependent accumulation, risk thresholds, and population prevalence, is partly shaped by measurement strategy. These findings underscore that analytic framework is an integral component of how somatic aging biomarkers are interpreted in large cohorts.

## Methods

### Study Population

The UK Biobank is a prospective population-based cohort comprising >500,000 participants aged 40–71 years at recruitment, conducted between 2006 and 2010. Participants were enrolled from across the United Kingdom and provided biological specimens, underwent physical assessments, and completed detailed questionnaires capturing demographic, lifestyle, and health-related information. Genomic DNA was extracted from whole blood, and genotyping was performed using high-density SNP arrays: 50,000 individuals were genotyped using the Affymetrix UK BiLEVE array, while the remaining samples were analyzed using the Affymetrix UK Biobank Axiom® array.

### mLOY Assessment in the UK Biobank using mLRRY

To investigate mLOY, we utilized SNP array intensity data, as described in^1, 2^. After excluding individuals who had withdrawn consent, we restricted our analysis to self-reported male participants (n = 223,402). Copy number variation on the Y chromosome was quantified using the Log R Ratio (LRR), which compares observed probe intensities to expected values. The median LRR was calculated across 691 probes located within the male-specific region of chromosome Y (MSY; chrY: 2,649,520–59,034,050, GRCh37/hg19), yielding a continuous metric of mLOY, referred to as mLRRY. Values near zero reflect normal Y chromosome dosage, while more negative values indicate increasing degrees of Y chromosome loss. Copy number estimates for chromosome X were similarly derived, generating mLRRX values based on X-specific probes. Individuals with atypical sex chromosome karyotypes other than mLOY (n = 428) were identified and excluded. A systematic bias between mLRRY and mLRRX was detected and corrected using linear regression, following established methods.^13^ For the final analytic cohort (n = 222,974), mLRRY values were converted to estimated percentages of leukocytes exhibiting mLOY using a validated transformation.^14^

### mLOY Assessment in the UK Biobank using MoChA

Raw genotyping array intensity data were processed to derive B-allele frequencies (BAF) and log2 R ratio (LRR) measures and analyzed using the Mosaic Chromosomal Alterations (MoChA) caller (v2025-08-19; https://github.com/freeseek/mochawdl) following the recommended MoChA workflow.^15^ MoChA applies a Viterbi-based hidden Markov model that jointly models phased BAF and LRR to detect mosaic chromosomal alterations. Genotype phasing was performed using SHAPEIT5 to infer haplotypes used by MoChA.

Quality control followed the standard MoChA filtering criteria: samples with call_rate < 0.97 or baf_auto > 0.03 were excluded. In addition, MoChA callsets were filtered according to the recommended MoChA call-level filters to remove low-confidence calls and likely germline events (including calls flagged as CNP and calls failing recommended LOD and event-prior thresholds).

mLOY was defined in male participants using the standard MoChA inference strategy based on allelic imbalance in the pseudoautosomal region 1 (PAR1). Specifically, males were classified as mLOY-positive when MoChA reported a chromosome X event (reflecting PAR1 analysis in male samples) with event length > 2 Mb and rel_cov < 2.5, which also helps exclude aneuploidies such as XXY and XYY. The mosaic cell fraction was estimated from BAF deviations within the PAR1 region, enabling sensitive detection of low-burden mLOY.

### Derivation of the mLRRY Based 8.81% mLOY Threshold in UK Biobank

To mitigate technical bias in mLRRY based estimates of mLOY, we applied a regression-based adjustment to account for array-wide intensity variation. This approach leverages the association between unadjusted mLRRY and mLRRX, with mLRRX serving as a proxy for global technical variation. Linear regression coefficients derived from this relationship were used to correct mLRRY values (Supplemental Figure 2A). The model was fitted in a filtered subset of samples (unadjusted mLRRY > −0.2), excluding influential observations based on Cook’s distance (<1.5×IQR above the third quartile; n = 198,389).

Following regression-based correction, a constant shift was applied to center the adjusted mLRRY distribution around zero for mLOY-negative samples. This constant was defined as the peak of the unadjusted mLRRY distribution and was estimated using kernel density estimation with the Sheather–Jones bandwidth method implemented in R. In this dataset, the estimated shift was 0.0018 (Supplemental Figure 2B).

LOY status was subsequently assigned using a threshold derived from the adjusted mLRRY distribution. Samples with adjusted mLRRY values below the 99% confidence limit of a simulated distribution representing technical mLRRY variation were classified as mLOY-positive (Supplemental Figure 2C; Supplemental Table 1). This threshold corresponds to an estimated mLOY fraction of 8.81% and was designed to minimize false-positive mLOY calls attributable to residual technical noise (Supplemental Figure 2D).

### Association of mLOY with all-cause mortality and clinical characteristics

We evaluated the association between %mLOY and all-cause mortality in male participants using three quantification methods (MoChA, mLRRY, and mLRRY [T = 8.81]). mLOY was categorized into predefined ranges (0%, >0–5%, >5–10%, >10–15%, and >15%), with individuals without detectable mLOY (0%) serving as the reference group. Time-to-event analyses were performed using Cox proportional hazards models with adjustment for age, body mass index (BMI), smoking status, alcohol consumption, and diabetes, and hazard ratios (HRs) with 95% confidence intervals were estimated for each mLOY category.

To further characterize dose-response relationships, mLOY was additionally modeled as a continuous variable using Cox regression with natural cubic splines. Analyses were restricted to individuals with detectable mLOY (>0%), with hazard ratios expressed relative to the no-mLOY reference category. Predicted HRs and 95% confidence intervals were derived across the observed range of mLOY to assess nonlinear associations.

Method-specific thresholds for elevated risk were defined using a data-driven approach based on spline models. The threshold was identified as the lowest mLOY value at which the lower bound of the 95% confidence interval for the hazard ratio exceeded unity. To ensure robustness, a conservative criterion was applied (lower bound >1.01). Using this approach, thresholds were estimated at 3.1% for MoChA and 10.1% for mLRRY. These empirically derived thresholds reflect inflection points in statistical risk and should not be interpreted as discrete biological cut-offs.

We further examined the distribution of demographic, lifestyle, and clinical characteristics among individuals exceeding these thresholds (MoChA ≥3.1%; mLRRY ≥10.1%). The prevalence of selected traits, including age >60 years, obesity (BMI >30), smoking, diabetes, hypertension, cancer, and incident cardiovascular outcomes (myocardial infarction, stroke, heart failure, and atrial fibrillation), was summarized as counts and proportions within each method.

### Computational Environment

All analyses were performed on a dedicated high-performance workstation running Ubuntu 24.04 LTS. The system was equipped with a multi-core Intel Xeon processor, 256 GB of RAM, and approximately 20 TB of local storage, providing sufficient memory and disk capacity to accommodate UK Biobank–scale genotyping array intensity files, MoChA intermediate outputs, and downstream statistical analyses.

### Study approval

The UK Biobank has approval from the North West Multi-centre Research Ethics Committee (covering the United Kingdom), the National Information Governance Board for Health & Social Care (covering 8 England and Wales) and the Community Health Index Advisory Group (for Scotland). A generic Research Tissue Bank approval granted by the National Research Ethics Service and UK Biobank’s governing Research Ethics Committee allows applicants to conduct research on UK Biobank data without obtaining separate ethical approvals. The UK Biobank dataset used in the present study received project ID 170256, all study procedures were performed in accordance with the ethical principles for medical research from the World Medical Association Declaration of Helsinki and all participants provided written informed consent.

## Data Availability

Not Applicable

