## Supplementary material for "Measurement strategy alters inferred age-dependent accumulation and mortality risk of mosaic Y loss": Supplemental Figures.pdf

**Supplemental Table 1.** Age-stratified prevalence of mLOY among male UK Biobank participants assessed using mLRRY filtered at a >8.81% cellular fraction threshold.

| | **mLRRY (> 8.81%)** |
|---|---|
| **Age** | **mLOY (Count %)** *(N=14,379)* |
| <41 | 12 (0.4) |
| 41-42 | 54 (0.5) |
| 43-44 | 60 (0.5) |
| 45-46 | 71 (0.6) |
| 47-48 | 86 (0.7) |
| 49-50 | 155 (1.2) |
| 51-52 | 208 (1.6) |
| 53-54 | 322 (2.4) |
| 55-56 | 479 (3.2) |
| 57-58 | 712 (4.5) |
| 59-60 | 1169 (6) |
| 61-62 | 1808 (7.9) |
| 63-64 | 2159 (10.4) |
| 65-66 | 2618 (12.9) |
| 67-68 | 2714 (16) |
| 69-73 | 1752(19.4) |

**mLOY**: mosaic loss of Y chromosome
**mLRRY**: median log-R ratio of the Y chromosome (pipeline)

### Supplementary Figure 1

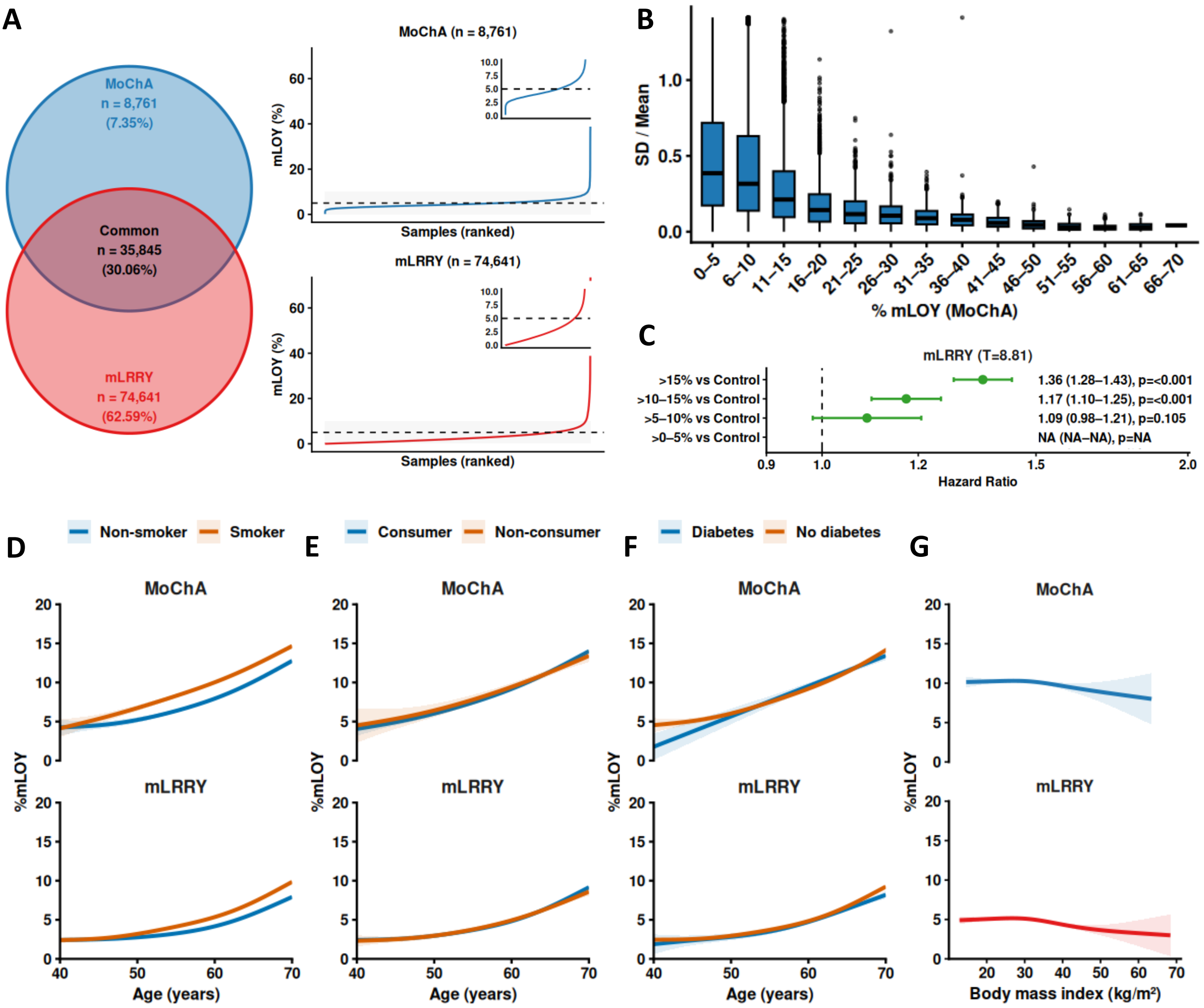


**Supplementary Figure 1. Concordance, variability, and demographic patterns of mLOY across methods.** **(A)** Overlap of individuals classified as mLOY-positive by MoChA and mLRRY. A total of 119,247 individuals were identified by at least one method, with 35,845 concordant calls. Rank-ordered distributions of mLOY% for method-specific calls (mLRRY-only and MoChA-only) show that discordant classifications are predominantly enriched at low mosaic fractions. Specifically, 63,889 (57.8%) of mLRRY-specific and 5,754 (12.9%) of MoChA-specific calls occur below 5% mLOY. **(B)** Inter-method variability in mLOY estimates across levels of mosaic burden. Boxplots show the coefficient of variation (SD/mean) between MoChA- and mLRRY-derived mLOY% estimates stratified by MoChA-defined mLOY bins. Variability is greatest at low mosaic fractions and decreases with increasing mLOY, indicating improved agreement at higher levels of mosaicism. **(C)** Association of mLOY categories with all-cause mortality. Hazard ratios (HRs) are estimated from Cox proportional hazards models comparing mLOY categories (>0-5%, >5-10%, >10-15%, >15%) to controls (0%), adjusted for age, body mass index, smoking status, alcohol consumption, and diabetes. Points indicate HRs and error bars denote 95% confidence intervals. **(D–G)** Age-dependent patterns of mLOY stratified by demographic and clinical factors. GAM spline curves show mLOY% as a function of age by (D) smoking status, (E) alcohol consumption, (F) diabetes status, and (G) BMI. Results are shown separately for MoChA (top panels) and mLRRY (bottom panels), with shaded areas representing 95% confidence intervals.

### Supplementary Figure 2

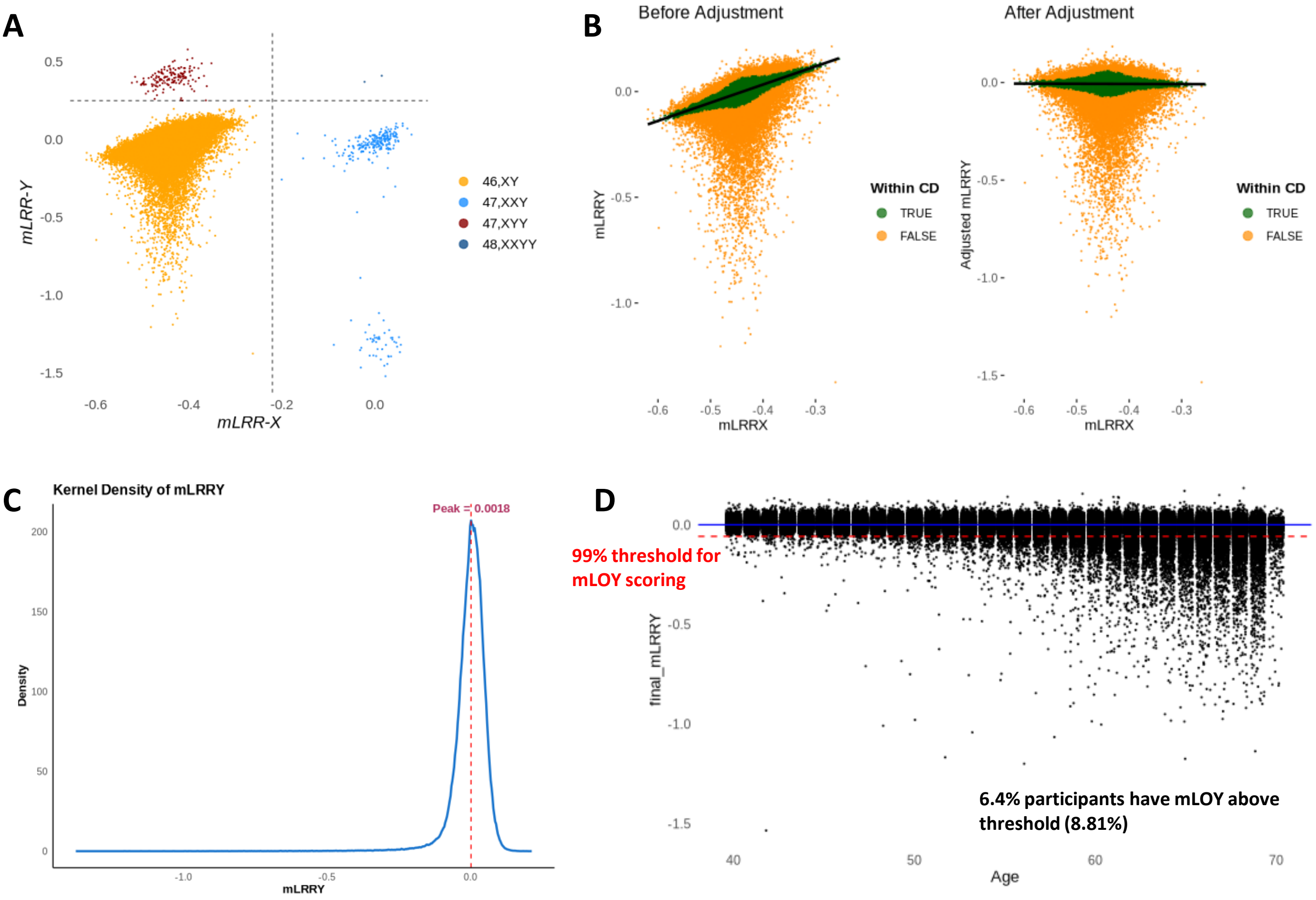


**Supplementary Figure 2. Derivation and calibration of mLRRY-based mosaic loss of chromosome Y (LOY) estimates from SNP array data in male UK Biobank participants. (A)** Dot plot of sex chromosome intensity measures showing mLRRY (Y-axis) and mLRRX (X-axis), calculated as the median Log R Ratio across Y- and X-chromosome probes, respectively. Thresholds of mLRRY < 0.25 and mLRRX < −0.22 were applied to exclude outliers (yellow; n = 222,974). **(B)** Correction of mLRRY values for technical variation using mLRRX as a proxy. Distributions of mLRRY values before and after regression-based adjustment are shown. **(C)** Centering of adjusted mLRRY values by applying a constant derived from the peak of the unadjusted mLRRY distribution. The constant (0.0018) was estimated using kernel density estimation in R with the Sheather–Jones ("SJ") bandwidth method. **(D)** Distribution of mosaic loss of chromosome Y (LOY) estimates in 222,974 male participants. The dashed line indicates the threshold corresponding to the 99th percentile of technical variation (mLRRY = −0.0579), which was used to classify individuals as LOY-positive or LOY-negative.
